# Severity of Respiratory Infections due to SARS-CoV-2 in Working Population: Age and Body Mass Index Outweigh ABO Blood Group

**DOI:** 10.1101/2020.11.05.20226100

**Authors:** Johannes Schetelig, Henning Baldauf, Sarah Wendler, Falk Heidenreich, Ruben Real, Martin Kolditz, Andrea Rosner, Alexander Dalpke, Katja de With, Vinzenz Lange, Jan Markert, Ralf Barth, Carolin Bunzel, Dennis Endert, Jan A Hofmann, Jürgen Sauter, Stefanie N Bernas, Alexander H Schmidt

## Abstract

**Background:** With increasing rates of SARS-CoV-2 infections and the intention to avoid a lock-down, the risks for the working population are of great interest. No large studies have been conducted which allow risk assessment for this population.

**Methods:** DKMS is a non-profit donor center for stem cell donation and reaches out to registered volunteers between 18 and 61 years of age. To identify risk factors for severe COVID-19 courses in this population we performed a cross-sectional study. Self-reported data on oro- or nasopharyngeal swabs, risk factors, symptoms and treatment were collected with a health questionnaire and linked to existing genetic data. We fitted multivariable logistic regression models for the risk of contracting SARS-CoV-2, risk of severe respiratory infection and risk of hospitalization.

**Findings:** Of 4,440,895 contacted volunteers 924,660 (20.8%) participated in the study. Among 157,544 participants tested, 7,948 reported SARS-CoV-2 detection. Of those, 947 participants (11.9%) reported an asymptomatic course, 5,014 (63.1%) mild/moderate respiratory infections, and 1,987 (25%) severe respiratory tract infections. In total, 286 participants (3.6%) were hospitalized for respiratory tract infections. The risk of hospitalization in comparison to a 20-year old person of normal weight was 2.1-fold higher (95%-CI, 1.2-3.69, p=0.01) for a person of same age with a BMI between 35-40 kg/m^2^, it was 5.33-fold higher (95%-CI, 2.92-9.70, p<0.001) for a 55-year old person with normal weight and 11.2-fold higher (95%-CI, 10.1-14.6, p<0.001) for a 55-year old person with a BMI between 35-40 kg/m^2^. Blood group A was associated with a 1.15-fold higher risk for contracting SARS-CoV-2 (95%-CI 1.08-1.22, p<0.001) than blood group O but did not impact COVID-19 severity.

**Interpretation:** In this relatively healthy population, the risk for hospitalizations due to SARS-CoV-2 infections was moderate. Age and BMI were major risk factors. These data may help to tailor risk-stratified preventive measures.

**Funding:** DKMS initiated and conducted this study. The Federal Ministry of Education and Research (BMBF) supported the study by a research grant (COVID-19 call (202), reference number 01KI20177).

## Introduction

With numbers of new SARS-CoV-2 infections reaching alarming levels many countries discuss and implement focused measures to reduce the spread of the virus and to avoid full lock-downs. The elderly and individuals who are at greater risk of COVID-19 related mortality due to comorbidity are in the center of this debate. Risk factor analyses from hospital-based or health insurance-based studies^1-5^ provide comprehensive data for these subpopulations. Yet, the working population may also face considerable risks, especially when lock-down measures and stay-at-home orders to reduce the spread of the virus are avoided. Owing to the design of most studies published so far, little information is available on risk factors for the working population, in spite of an increasing share of younger COVID-19 patients in the second half of 2020^6-7^.

DKMS gemeinnützige GmbH (DKMS) is a major stem cell donor registry and reaches out to more than ten million volunteers who registered as potential stem cell donor^8^. Volunteers may only register if their general health condition does not preclude them from a stem cell donation. Registered donors are requested to report relevant medical information, which may affect their suitability as stem cell donors and may subsequently be excluded from the active donor file. DKMS excludes volunteers from the registry at the age of 61 years. As a consequence, volunteers registered with DKMS represent a subset of the working population with slightly less health issues^9^.

Since the severity of SARS-CoV-2 infections varies from asymptomatic courses to acute respiratory failure, information on risk factors is critical. Besides age, comorbidity, and risk behaviour, genetic data may partly explain this variation. As a donor registry, DKMS administrates immunogenetic data relevant for donor-patient matching in the context of stem cell transplantation. In response to the pandemic, DKMS therefore launched a large population-based study to identify risk factors, which might help individual risk assessment and tailor preventive measures.

Blood group A has been associated with an increased risk of contracting the virus and/or severe courses^10-13^. While the pathophysiology is unclear, it was speculated that A epitopes facilitate virus entry. ABO epitopes modify many proteins, lipids and sphingolipids. Individuals with at least one *A*_*1*_ allele have a higher glycosyltransferase activity and thus ubiquitously expose much higher numbers of A epitopes compared to individuals who inherited *A*_*2*_ alleles but no *A*_*1*_ allele^14^. Therefore, we tested the hypothesis that individuals with A_1_ phenotype have a greater risk of infection compared to individuals with A_2_ phenotype.

Here, we present first results of this immunogenetic study. Age and body mass index (BMI) were major risk factors for severe respiratory tract infections and hospitalization for respiratory distress. In our data, ABO blood group (BG) was a minor factor for the risk of infection but not for COVID-19 severity. Comparable rates of infection among participants with BG A_1_ versus A_2_, as observed in our study, strongly argue against the hypothesis that A epitopes facilitate virus entry into host cells.

## Methods

### Study design

The project was designed as a registry-based cross-sectional study. Existing immunogenetic data were linked to self-reported COVID-19 specific data collected with a standardized health questionnaire. The responsible Institutional Review Board of the Technische Universität Dresden (IRB00001473) approved the study. We registered the study with the trial registry of the German Center for Infection Research (https://dzif.clinicalsite.org/de/cat/2099/trial/4361). Data privacy of the participating individuals was protected in accordance with the General Data Protection Regulation of the European Union. We conducted the study in compliance with the principles of the Declaration of Helsinki. All participants provided explicit consent that COVID-19 specific data were linked to immunogenetic data in the DKMS donor registry file.

### Study population

Study invitations were sent by e-mail to volunteers registered with DKMS in Germany who were between 18 and 61 years old, had previously provided an e-mail address, and had not objected to being contacted for issues other than stem cell donation requests. At registration with DKMS, volunteers provide basic health information with respect to weight, height, and serious health conditions. As a consequence, volunteers registered with DKMS represent a subset of the working population characterized by the exclusion of individuals with serious chronic illnesses at the time of registration. Volunteers who develop serious acute or chronic health conditions stay in the registry unless they proactively contact DKMS or get contacted for a stem cell donation request.

### Health questionnaire

In total, 4,440,895 donors were contacted via e-mail for study participation between August 17^th^, 2020 and August 26^th^, 2020. Participants who provided informed consent completed the health questionnaire through a web-based password protected log-in. The database was locked on September 14^th^, 2020. The brief health questionnaire contained questions on the testing procedure, known risk factors, COVID-19 self-reported symptoms, and the severity of the course indicated by questions about hospitalization, need for supplemental oxygen, and mechanical ventilation. A translated version of the health questionnaire is provided in the supplement (Supplemental Material 1).

### Data definitions and processing

#### Case definitions

Participants who reported at least one positive test for SARS-CoV-2 on an oro- or nasopharyngeal swab were considered as positive cases. Participants who had been tested at least once but never tested positive for SARS-CoV-2 were assigned as negative cases.

#### Phenotype definitions

Based on self-reported symptoms and treatment information, participants who reported a positive test were classified in three groups: i) severe respiratory tract infections (RTI) were defined by the combination of at least fever and cough, dyspnea and cough, dyspnea and fever, or dyspnea and myalgia^5^. This group included respiratory hospitalizations, defined by in-patient care with supplemental oxygen or mechanical ventilation or hospitalization for dyspnea or cough; ii) mild/moderate courses, if self-reported symptoms did not meet the definition of severe RTI. These participants reported any other of the following symptoms fever, myalgia, sore throat, loss of taste/smell, cough, or dyspnea; and iii) participants without symptoms.

#### Definition of risk factors

Obesity was classified into categories as suggested by the Centers of Disease Control and Prevention (https://www.cdc.gov/obesity/adult/defining.html). A history of arterial hypertension and diabetes mellitus was considered only when medication to treat this condition was reported for 2019.

### Immunogenetic data

Volunteers who registered with DKMS were typed upfront with a panel of genes potentially relevant for donor selection. This panel evolved over years and includes ABO and Rh blood groups. Genotyping of DNA extracted from buccal swabs is performed at high resolution with an amplicon-based approach using Illumina devices^15-16^.

Based on information for *ABO* alleles, heterozygous alleles were grouped into ABO BG phenotypes by assuming dominant expression for *A*_*1*_ over *A*_*2*_ and BG A or B over BG O. Blood group A_1_ was coded for individuals with *A*_*1*_*+0, A*_*1*_*+A*_*1*_, *A*_*1*_*+A*_*2*_, *A*_*1*_*+A*_*3*_, *A*_*1*_*+A*_*w*_, *or A*_*1*_*+A*_*x*_ and BG A_2_ was coded for individuals with *A*_*2*_*+0, A*_*2*_*+A*_*2*_, *A*_*2*_*+A*_*x*_, *or A*_*2*_*+A*_*el*_. Ambiguous genotypes were imputed by the most frequent allele according to Lang et al 2016 ^16^.

### Statistical methods

#### Analysis populations

We defined two analysis populations: 1) The population to analyze the risk of contracting SARS-CoV-2 comprised all positive and negative cases. 2) The population to analyze risk factors for COVID-19 severity included all positive cases but not individuals with positive tests in August or September, because they might not have experienced the maximum severity of COVID-19 at the time of reporting. Details on participant withdrawals and exclusions and on population sizes are provided in supplemental Figure S1.

#### Statistical analysis

In the descriptive part we used frequencies and percentages to describe the data. The risk of contracting SARS-CoV-2 was specified as primary endpoint for the immunogenetic evaluation of the impact of ABO blood groups. Five *a priori* defined hypotheses (A_1_ vs A_2_, A vs AB vs B vs O, A vs non-A, O vs non-O, and AB vs non-AB) were tested. For each single test a one-sided significance level of 1% was set in order to control of type I error. The power for these tests exceeded 96%. Binary logistic regression models were used to investigate potential factors associated with the risk of infection and the severity of COVID-19. Odds ratios (ORs) and their 95% confidence intervals were used to describe the associations. For exploratory and confirmatory analyses, statistical testing was based on Wald tests for logistic regression coefficients and likelihood ratio tests. All models were adjusted for age, sex, BMI, diabetes mellitus, arterial hypertension, and smoking status. Age, BMI, and month of positive test were modeled as categorical variables. All analyses were carried out using R Statistical Software version 3.5.1.

## Results

### Study participation

In total 4,440,895 e-mails were sent and 924,660 participants consented to study participation (20.8% return rate). Response rates of female and male email recipients were 21.8% and 16.5% (below 40 years) and 24.5% and 20.0% (above 40 years), respectively (chi-square test, p<0.001). Regional response rates ranged between 17% and 25%.

### Descriptive analysis

Altogether 157,544 participants reported known results from a test for SARS-CoV-2 and 7,948 participants reported a positive test. Heat maps for regional rates of positive tests reported in this survey mirrored regional incidences of SARS-CoV-2 infections reported to the Robert Koch Institute (RKI) (Supplemental Figure 2). According to this source, the cumulative incidence of confirmed SARS-CoV-2 infections until August 31^st^ among individuals aged between 15 and 59 years ranged between 0.33% and 0.38%. The overall rate of SARS-CoV-2 positive participants among all study participants was 0.86% (7,948/924,561).

We show demographic information of SARS-CoV-2 positive and -negative participants in Table 1. Female predominance among study participants also reflects the fact that more women than men are registered with DKMS (61% vs. 39%, respectively). Among participants who tested SARS-CoV-2 positive, 947 individuals (11.9%) reported no symptoms related to the infection, 5,014 individuals (63.1%) reported mild/moderate symptoms and 1,987 individuals (25.0%) reported symptoms of severe RTI. Respiratory hospitalization was reported from 286 participants (3.6% out of all SARS-CoV-2 positive individuals). Of those, 161 participants (2.0%) needed supplemental oxygen and 22 participants (0.28%) needed mechanical ventilation. Participants who reported respiratory support had a median age of 50 years (range: 24 to 58 years). Twenty participants were hospitalized, but did not meet the criteria for respiratory hospitalization.

**Table 1.**
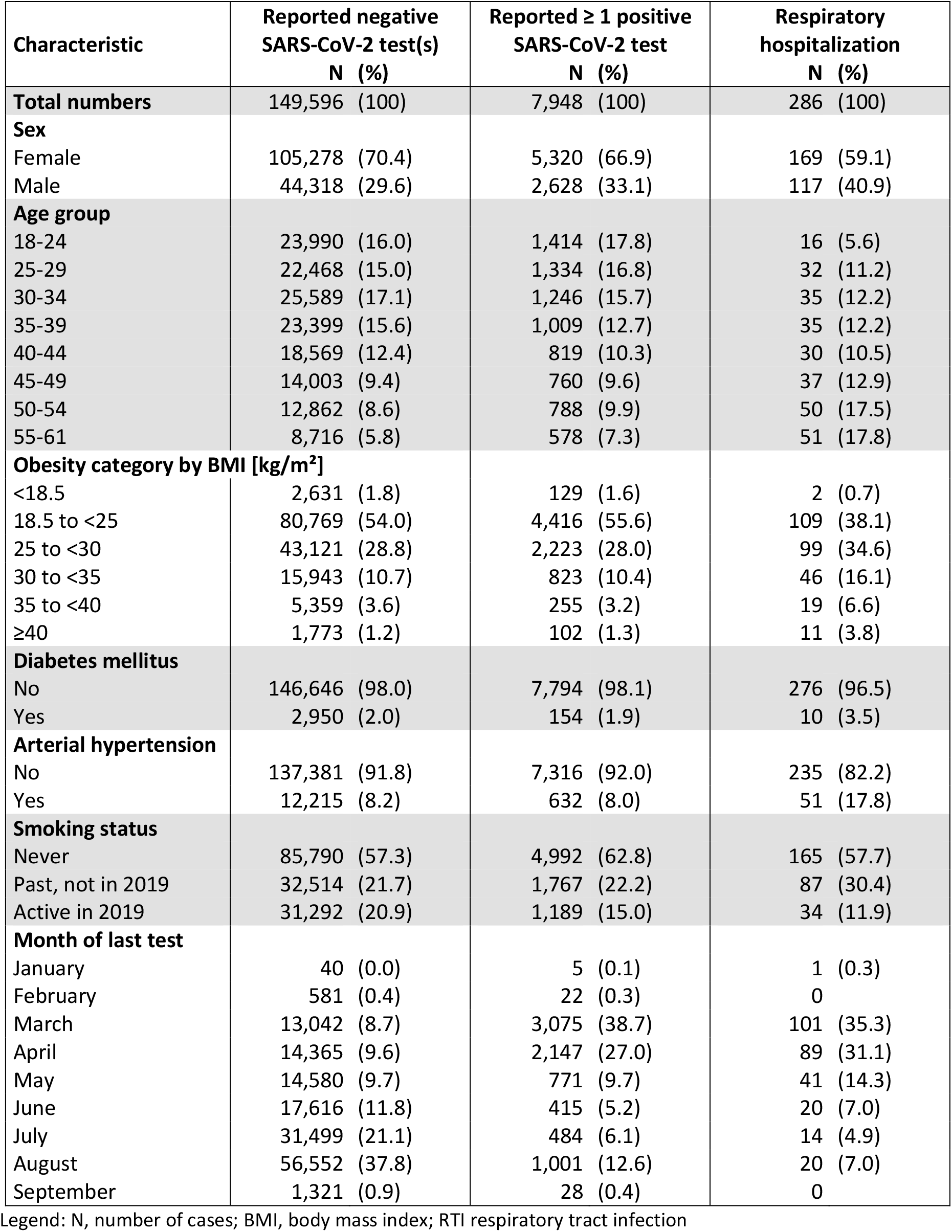
Characteristics of SARS-CoV-2 positive and negative participants.

| Characteristic | Reported negative<br>SARS-CoV-2 test(s)<br>N (%) | Reported ≥ 1 positive<br>SARS-CoV-2 test<br>N (%) | Respiratory<br>hospitalization<br>N (%) |
| --- | --- | --- | --- |
| **Total numbers** | 149,596 (100) | 7,948 (100) | 286 (100) |
| **Sex** |  |  |  |
| Female | 105,278 (70.4) | 5,320 (66.9) | 169 (59.1) |
| Male | 44,318 (29.6) | 2,628 (33.1) | 117 (40.9) |
| **Age group** |  |  |  |
| 18-24 | 23,990 (16.0) | 1,414 (17.8) | 16 (5.6) |
| 25-29 | 22,468 (15.0) | 1,334 (16.8) | 32 (11.2) |
| 30-34 | 25,589 (17.1) | 1,246 (15.7) | 35 (12.2) |
| 35-39 | 23,399 (15.6) | 1,009 (12.7) | 35 (12.2) |
| 40-44 | 18,569 (12.4) | 819 (10.3) | 30 (10.5) |
| 45-49 | 14,003 (9.4) | 760 (9.6) | 37 (12.9) |
| 50-54 | 12,862 (8.6) | 788 (9.9) | 50 (17.5) |
| 55-61 | 8,716 (5.8) | 578 (7.3) | 51 (17.8) |
| **Obesity category by BMI [kg/m<sup>2</sup>]** |  |  |  |
| <18.5 | 2,631 (1.8) | 129 (1.6) | 2 (0.7) |
| 18.5 to <25 | 80,769 (54.0) | 4,416 (55.6) | 109 (38.1) |
| 25 to <30 | 43,121 (28.8) | 2,223 (28.0) | 99 (34.6) |
| 30 to <35 | 15,943 (10.7) | 823 (10.4) | 46 (16.1) |
| 35 to <40 | 5,359 (3.6) | 255 (3.2) | 19 (6.6) |
| ≥40 | 1,773 (1.2) | 102 (1.3) | 11 (3.8) |
| **Diabetes mellitus** |  |  |  |
| No | 146,646 (98.0) | 7,794 (98.1) | 276 (96.5) |
| Yes | 2,950 (2.0) | 154 (1.9) | 10 (3.5) |
| **Arterial hypertension** |  |  |  |
| No | 137,381 (91.8) | 7,316 (92.0) | 235 (82.2) |
| Yes | 12,215 (8.2) | 632 (8.0) | 51 (17.8) |
| **Smoking status** |  |  |  |
| Never | 85,790 (57.3) | 4,992 (62.8) | 165 (57.7) |
| Past, not in 2019 | 32,514 (21.7) | 1,767 (22.2) | 87 (30.4) |
| Active in 2019 | 31,292 (20.9) | 1,189 (15.0) | 34 (11.9) |
| **Month of last test** |  |  |  |
| January | 40 (0.0) | 5 (0.1) | 1 (0.3) |
| February | 581 (0.4) | 22 (0.3) | 0 |
| March | 13,042 (8.7) | 3,075 (38.7) | 101 (35.3) |
| April | 14,365 (9.6) | 2,147 (27.0) | 89 (31.1) |
| May | 14,580 (9.7) | 771 (9.7) | 41 (14.3) |
| June | 17,616 (11.8) | 415 (5.2) | 20 (7.0) |
| July | 31,499 (21.1) | 484 (6.1) | 14 (4.9) |
| August | 56,552 (37.8) | 1,001 (12.6) | 20 (7.0) |
| September | 1,321 (0.9) | 28 (0.4) | 0 |

Legend: N, number of cases; BMI, body mass index; RTI respiratory tract infection

### Rates of positive tests

We assessed risk factors for contracting SARS-CoV-2 by comparing the distribution of baseline characteristics between participants who tested positive and those who reported a negative test in the same period. The rate of positive tests by age group showed a bimodal distribution with peak rates among individuals aged 25 to 29 years (5.6%) and 50 to 61 years (6.2%). It was lowest among participants aged 35 to 39 years (4.1%) (see Figure 1). Men (5.6%) reported higher rates of positive tests compared to women (4.6%) (adjusted OR 1.27, 95%-CI: 1.21-1.34; p<0.001). Active smokers (3.7%) reported lower rates of positive tests than non-smokers (5.5%) (adjusted OR 0.63, 95%-CI: 0.59-0.68; p<0.001). Results of multivariable logistic regression models are shown in Table 2.

**Table 2.** Multivariable risk factor analysis.

| Characteristic | Contracting SARS-CoV-2 |  |  | Severe respiratory tract infection |  |  | Respiratory hospitalization |  |  |
| --- | --- | --- | --- | --- | --- | --- | --- | --- | --- |
|  | OR | (95%-CI) | p | OR | (95%-CI) | p | OR | (95%-CI) | p |
| **Sex** |  |  |  |  |  |  |  |  |  |
| Female | 1 |  |  | 1 |  |  | 1 |  |  |
| Male | 1.27 | (1.21-1.34) | <.001 | 0.64 | (0.57-0.73) | <.001 | 1.22 | (0.94-1.59) | 0.133 |
| **Age group** |  |  |  |  |  |  |  |  |  |
| 18-24 | 1 |  |  | 1 |  |  | 1 |  |  |
| 25-29 | 1.00 | (0.93-1.09) | 0.931 | 1.13 | (0.93-1.38) | 0.214 | 1.75 | (0.94-3.25) | 0.075 |
| 30-34 | 0.82 | (0.76-0.89) | <.001 | 1.15 | (0.94-1.40) | 0.189 | 1.84 | (1.00-3.40) | 0.051 |
| 35-39 | 0.74 | (0.68-0.81) | <.001 | 1.36 | (1.11-1.68) | 0.004 | 2.28 | (1.23-4.20) | 0.009 |
| 40-44 | 0.74 | (0.67-0.81) | <.001 | 1.36 | (1.09-1.69) | 0.007 | 2.24 | (1.18-4.23) | 0.013 |
| 45-49 | 0.89 | (0.81-0.98) | 0.019 | 1.42 | (1.14-1.78) | 0.002 | 3.16 | (1.72-5.80) | <.001 |
| 50-54 | 0.98 | (0.88-1.07) | 0.540 | 1.45 | (1.16-1.82) | 0.001 | 3.84 | (2.12-6.94) | <.001 |
| 55-61 | 1.04 | (0.94-1.16) | 0.455 | 1.39 | (1.08-1.78) | 0.009 | 5.33 | (2.92-9.70) | <.001 |
| **Obesity category by BMI [kg/m<sup>2</sup>]** |  |  |  |  |  |  |  |  |  |
| <18.5 | 0.91 | (0.76-1.10) | 0.319 | 0.78 | (0.47-1.28) | 0.32 | 0.43 | (0.06-3.14) | 0.407 |
| 18.5 to <25 | 1 |  |  | 1 |  |  | 1 |  |  |
| 25 to <30 | 0.91 | (0.86-0.96) | 0.001 | 1.27 | (1.12-1.45) | <.001 | 1.35 | (1.00-1.83) | 0.048 |
| 30 to <35 | 0.94 | (0.87-1.02) | 0.126 | 1.66 | (1.40-1.98) | <.001 | 1.66 | (1.14-2.41) | 0.009 |
| 35 to <40 | 0.88 | (0.77-1.01) | 0.069 | 2.02 | (1.51-2.68) | <.001 | 2.10 | (1.20-3.69) | 0.010 |
| ≥40 | 1.09 | (0.88-1.35) | 0.406 | 2.46 | (1.60-3.78) | <.001 | 3.24 | (1.56-6.74) | 0.002 |
| **Diabetes mellitus** |  |  |  |  |  |  |  |  |  |
| No | 1 |  |  | 1 |  |  | 1 |  |  |
| Yes | 1.03 | (0.86-1.22) | 0.763 | 1.59 | (1.10-2.28) | 0.012 | 1.62 | (0.82-3.20) | 0.161 |
| **Arterial hypertension** |  |  |  |  |  |  |  |  |  |
| No | 1 |  |  | 1 |  |  | 1 |  |  |
| Yes | 0.91 | (0.83-1.00) | 0.051 | 0.98 | (0.80-1.20) | 0.857 | 1.26 | (0.87-1.81) | 0.221 |
| **Smoking status** |  |  |  |  |  |  |  |  |  |
| Never | 1 |  |  | 1 |  |  | 1 |  |  |
| Past, not in 2019 | 0.96 | (0.91-1.02) | 0.250 | 1.10 | (0.96-1.25) | 0.180 | 1.12 | (0.84-1.49) | 0.452 |
| Active in 2019 | 0.63 | (0.59-0.68) | <.001 | 0.82 | (0.69-0.96) | 0.015 | 0.88 | (0.59-1.31) | 0.533 |
| **Month of last test** |  |  |  |  |  |  |  |  |  |
| January/February | 0.29 | (0.20-0.42) | <.001 | 1.22 | (0.53-2.84) | 0.639 | 1.02 | (0.14-7.69) | 0.986 |
| March | 1.54 | (1.45-1.64) | <.001 | 0.99 | (0.87-1.12) | 0.883 | 0.76 | (0.56-1.01) | 0.062 |
| April | 1 |  |  | 1 |  |  | 1 |  |  |
| May | 0.36 | (0.32-0.39) | <.001 | 1.18 | (0.98-1.42) | 0.077 | 1.29 | (0.88-1.90) | 0.192 |
| June | 0.16 | (0.14-0.18) | <.001 | 0.88 | (0.68-1.12) | 0.292 | 1.20 | (0.72-1.99) | 0.478 |
| July | 0.10 | (0.09-0.11) | <.001 | 0.84 | (0.66-1.06) | 0.144 | 0.71 | (0.40-1.26) | 0.240 |
| August | 0.11 | (0.10-0.12) | <.001 | excluded |  |  | excluded |  |  |
| September | 0.14 | (0.09-0.20) | <.001 | excluded |  |  | excluded |  |  |

Legend: OR, odds ratio; CI, confidence interval; p, p value; BMI, body mass index; RTI, respiratory tract infection. Reference categories are indicated by 1. Odds ratios were calculated in a multivariable logistic regression model containing information on age, sex, BMI, diabetes mellitus, arterial hypertension and smoking status.

**Figure 1.**
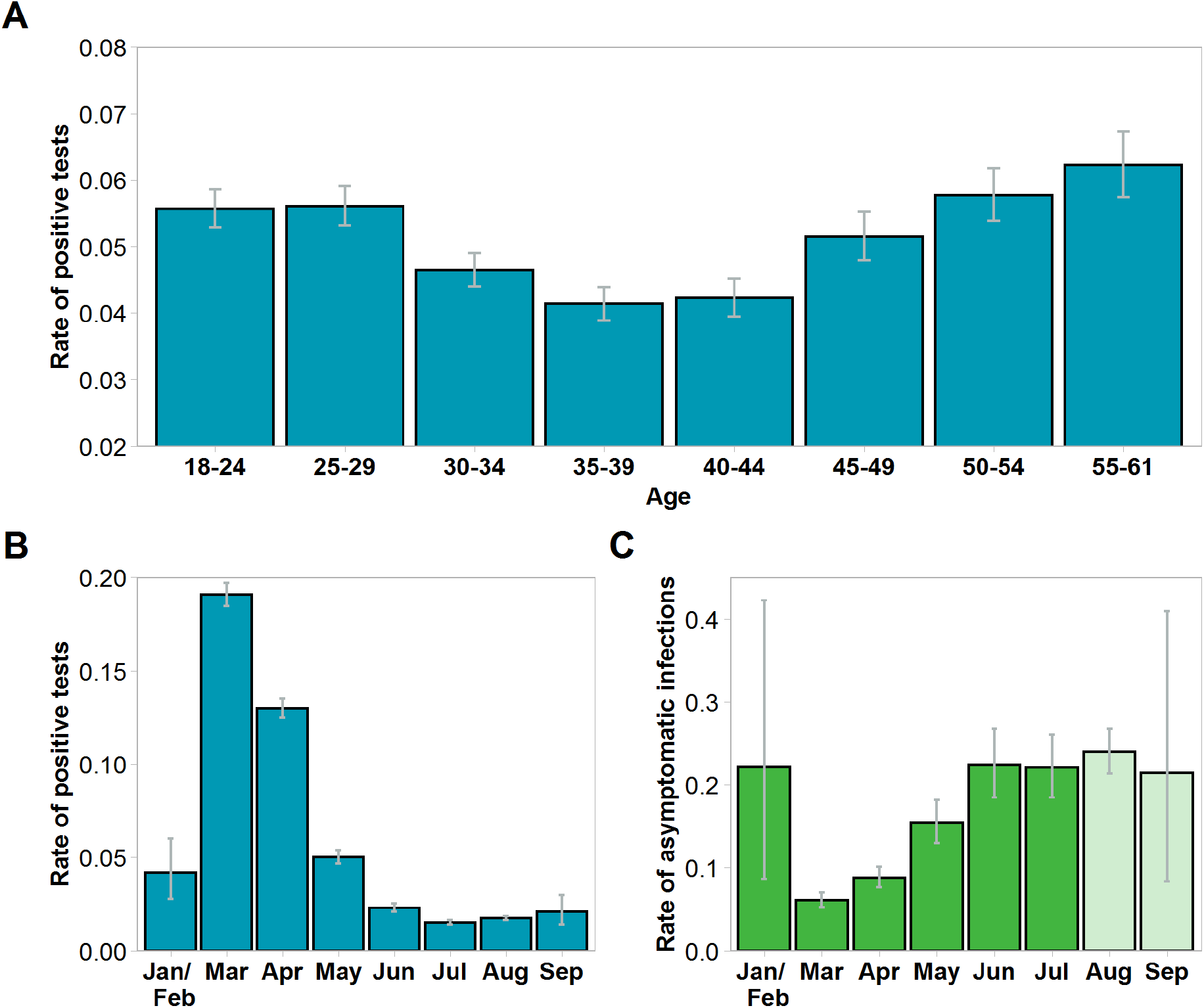
Percentage of positive tests for SARS-CoV-2 by age and month and rate of asymptomatic infections by month. Panel A shows the percentage of SARS-CoV-2 positive tests by age group. Panel B shows the percentage of positive tests by month. Nationwide percentages of positive tests as reported to the Robert Koch Institute (RKI) were 3% in January/February, 7% in March, 8% in April, 2% in May and 1% each from June to September but may include multiple tests per patient^17^. Panel C shows the percentage of participants who were tested positive and reported asymptomatic infections. Data for August and September (light green color) must be interpreted with caution as asymptomatic infections reported in these months might have been reported by individuals who had not yet observed symptoms at the time of reporting.

**Figure 2.**
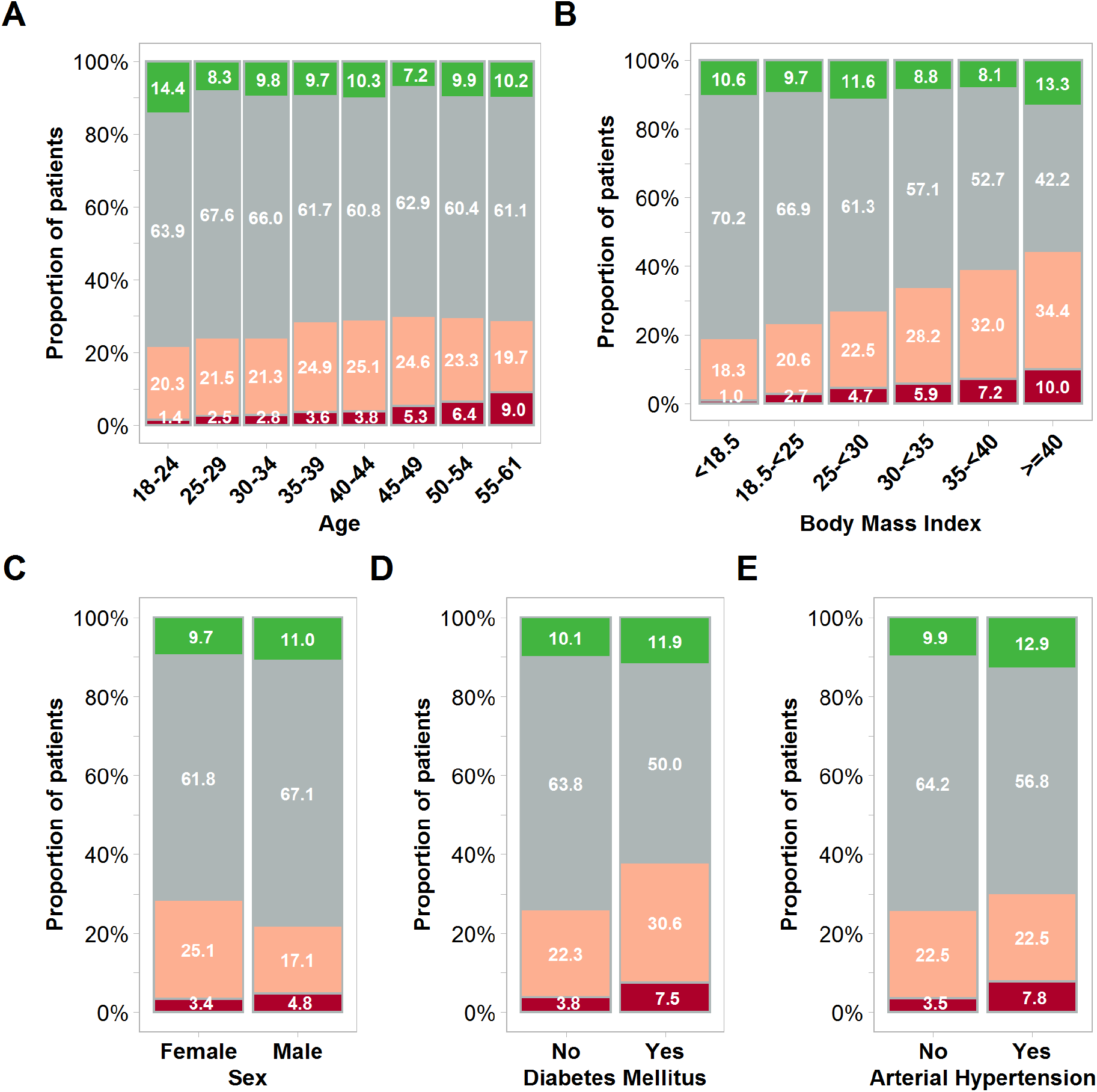
Severity of SARS-CoV-2 infections by age, BMI, sex, and medical history. The severity of self-reported COVID-19 courses is shown for all individuals who reported a positive test for SARS-CoV-2 from an oro- or nasopharyngeal swab between January and July. Results are shown by age group (Panel A), body mass index (Panel B), sex (Panel C), diabetes medication (Panel D), and arterial hypertension medication (Panel E). Red color indicates respiratory hospitalizations, defined by in-patient care with supplemental oxygen or mechanical ventilation or hospitalization for dyspnea or cough. Rose color indicates severe respiratory tract infections defined by the combination of at least fever and cough, dyspnea and cough, dyspnea and fever, or dyspnea and myalgia, but no need for hospitalization. Gray color represents mild/moderate courses. These participants did not meet the criteria for severe respiratory tract infections but reported symptoms such as fever, myalgia, sore throat, loss of taste/smell, cough, or dyspnea.

Rates of positive cases were also associated with the month of testing (Figure 1, Panel B and C). The rates of positive cases declined from 19.1% in March to 1.7% in August in our study. In parallel, the percentage of positive tests in Germany reported to the RKI peaked in late March/early April and declined in the subsequent four months until September 2020^17^.

### Risk factors contributing to COVID-19 severity

Age, BMI, and a history of diabetes or arterial hypertension had a significant impact on severe RTI and respiratory hospitalizations (Figure 2). Increasing age and BMI were associated with increased rates of respiratory hospitalizations due to SARS-CoV-2. Participants aged 55 to 61 years had a 5.3-fold greater risk (95%-CI: 2.9-9.7; p<0.001) for respiratory hospitalizations compared to participants aged 18 to 24 years. Participants with class 3 obesity (BMI ≥ 40) had a 3.2-fold greater risk (95%-CI: 1.6 to 6.7; p=0.002) of respiratory hospitalizations compared to individuals with normal weight. Tests for interactions between sex and BMI were not significant at the 5% level. According to the multivariable regression model (Table 2) the risk of respiratory hospitalization, for example for a 55-year old individual with a BMI of 35-40 kg/m^2^ was 11.2-fold higher compared to a 20-year old individual with normal weight (observed rates were 29% and 1.3%, respectively).

The risks for respiratory hospitalizations for participants with a history of diabetes or arterial hypertension were reflected by odds ratios of 1.62 (95%-CI: 0.82-3.20, p=0.16) and 1.26 (95%-CI: 0.87-1.81, p=0.22) with 95%-confidence intervals including 1, respectively. Past smokers and active smokers did not show an increased risk of respiratory hospitalizations (odds ratios 1.12, 95%-CI: 0.84-1.49, p=0.45 and 0.88, 95%-CI 0.59-1.31, p=0.53, respectively) compared to non-smokers.

Broad testing of asymptomatic individuals and contact persons of infected individuals started in Germany only in summer 2020. The rate of asymptomatic infections correlated with cases reported later in the year and with a lower percentage of positive tests. Participants aged 24 years or lower showed the highest incidence of asymptomatic infections (14.4%), but with large differences and a clear trend, e.g. 26.8%, 20.6% and 10.4% for individuals aged 18, 19, and 24 years, respectively.

### Impact of ABO/Rhesus phenotypes and genotypes

The impact of *ABO* genotype was analyzed in 125,125 participants with *ABO* genotypes, including 102,426 participants with allele-level resolution. We hypothesized that epitope density of ABO antigens is associated with the risk of infection. Participants with A_1_ and A_2_ phenotypes, however, had comparable rates of positive tests (5.3% and 5.4%, adjusted OR 1.03, 95%-CI 0.93-1.15; p=0.58) (see Figure 3A). Next, we analyzed the impact of ABO phenotypes on the risk of contracting SARS-CoV-2 and the risk of severe RTI (Figure 3A-D). Participants with BG A had a significantly higher rate of positive tests (adjusted OR 1.15, 95%-CI 1.08-1.22; p<0.001) compared to participants with BG O (see Figure 3C). Participants with BG AB (adjusted OR 1.19, 95%-CI 1.06-1.35; p=0.004) had significantly more infections as well, but no difference was found for participants with BG B compared to BG O.

**Figure 3.**
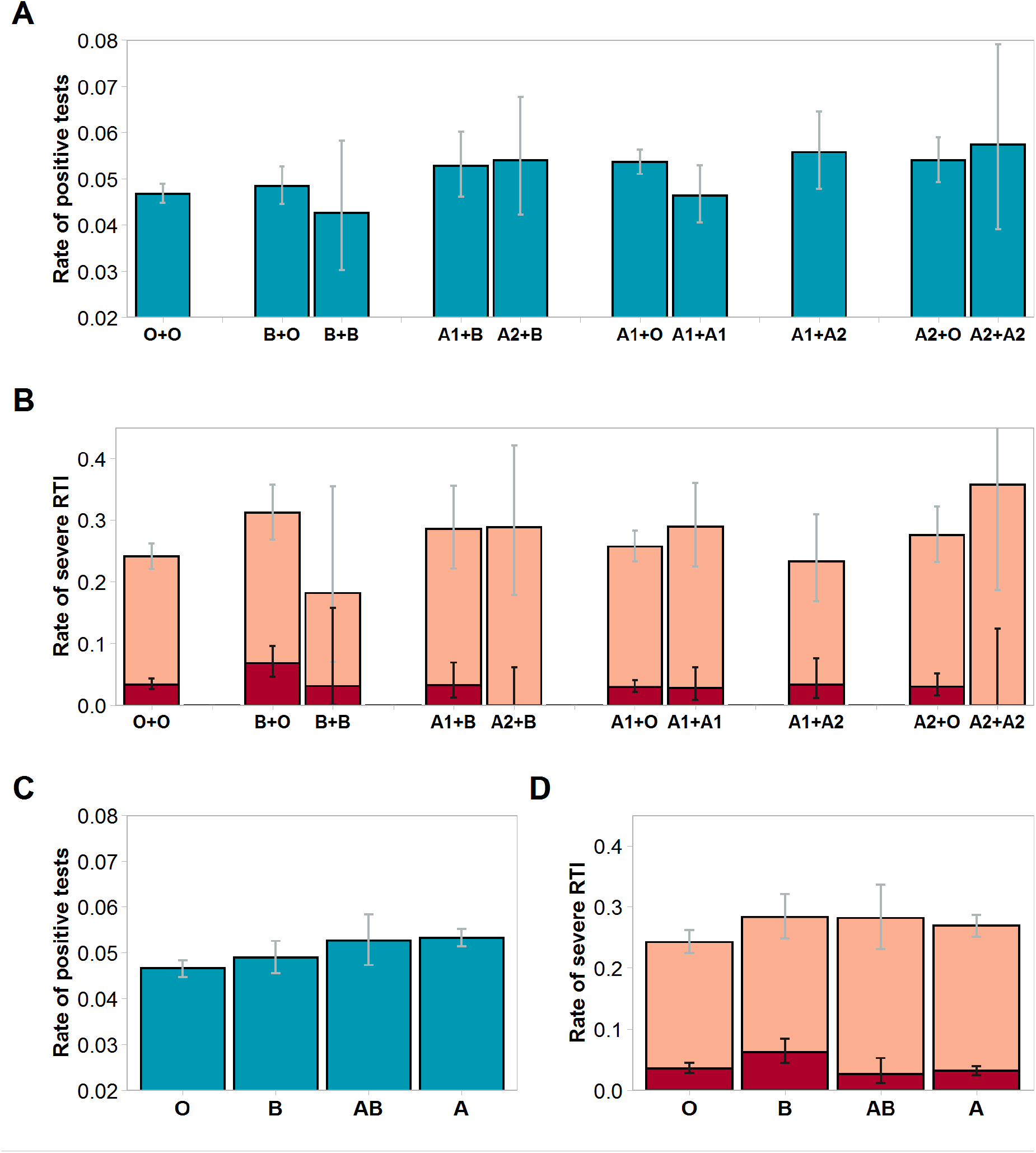
Rates of positive SARS-CoV-2 tests and severe respiratory tract infection by *ABO* blood group genotype and phenotype. Rates of positive tests (blue bars) by *ABO* genotype and phenotype are shown in panels A and C, respectively. Rates of severe respiratory tract infections and respiratory hospitalizations by *ABO* genotype and phenotype are shown in panels B and D. Respiratory hospitalization (red bars) was defined by in-patient care with supplemental oxygen or mechanical ventilation or hospitalization for dyspnea or cough. Severe respiratory tract infection (rose bar) was defined by the combination of at least fever and cough, dyspnea and cough, dyspnea and fever, or dyspnea and myalgia, but no need for hospitalization. Eighty-four were excluded from the comparisons displayed in panels A/C and five participants from the comparisons displayed in panels B/D, respectively, because they had rare genotypes, which could not be assigned to the 10 genotypes which are displayed.

In contrast, BG A did not show an impact on COVID-19 severity. But BG B was associated with a higher risk of severe RTI (OR 1.24, 95%-CI 1.01-1.53; p=0.038) and respiratory hospitalizations (OR 1.78, 95%-CI 1.18-2.68; p=0.006) compared to BG O (Figures 3B and 3D).

Finally, we performed exploratory analyses on the impact of *ABO* genotypes (Table 3). Allele level information was available for 102,426 participants. Participants who were homozygous for *B* had the lowest percentage of positive tests and participants homozygous for *A*_*2*_ had the highest percentage of positive tests. The adjusted odds ratio (OR 1.07, 95%-CI 0.96-1.19; p=0.23) for a positive test for participants with *A*_*2*_ alleles (*A*_*2*_+*A*_*2*_, *A*_*2*_+*O, A*_*2*_+*B*) versus *A*_*1*_ alleles (*A*_*1*_+*A*_*1*_, *A*_*1*_+*O, A*_*1*_+*B*) was not statistically significant. We provide additional information on exploratory analyses for blood group subtypes and rare genotypes in the supplement. Rhesus positivity was not associated with either phenotype in our study (Supplemental Table S1).

**Table 3.** Impact of *ABO* genotypes and phenotypes.

| Geno-<br>type | Contracting SARS-CoV-2 |  |  |  | Evaluable*<br>infections<br>N | Severe respiratory tract<br>infections |  |  |  | Respiratory hospitalizations |  |  |
| --- | --- | --- | --- | --- | --- | --- | --- | --- | --- | --- | --- | --- |
|  | Tests (%) | Cases | OR (95%-CI) | p |  | N | OR (95%-CI) | p |  | N | OR (95%-CI) | p |
| *O + O* | 40,975 (40.0) | 1,916 | 1 |  | 1,653 | 398 | 1 |  |  | 55 | 1 |  |
| *B + O* | 10,997 (10.7) | 533 | 1.05 (0.94-1.16) | 0.392 | 443 | 138 | 1.45 (1.14-1.83) | 0.002 |  | 30 | 2.16 (1.36-3.45) | 0.001 |
| *B + B* | 869 (0.8) | 37 | 0.95 (0.68-1.34) | 0.784 | 33 | 6 | 0.71 (0.29-1.77) | 0.467 |  | 1 | 0.79 (0.10-6.07) | 0.817 |
| *B + A<sub>1</sub>* | 4,072 (4.0) | 215 | 1.17 (1.01-1.36) | 0.042 | 186 | 53 | 1.29 (0.92-1.83) | 0.140 |  | 6 | 1.01 (0.42-2.39) | 0.987 |
| *B + A<sub>2</sub>* | 1,298 (1.3) | 70 | 1.29 (1.00-1.66) | 0.052 | 59 | 17 | 1.30 (0.73-2.33) | 0.377 |  | 0 | 0 | - |
| *A<sub>1</sub> + O* | 27,251 (26.6) | 1,462 | 1.15 (1.07-1.23) | <.001 | 1229 | 316 | 1.09 (0.92-1.30) | 0.311 |  | 36 | 0.89 (0.58-1.37) | 0.589 |
| *A<sub>1</sub> + A<sub>1</sub>* | 4,612 (4.5) | 214 | 0.99 (0.85-1.15) | 0.860 | 187 | 54 | 1.26 (0.90-1.77) | 0.180 |  | 5 | 0.78 (0.31-1.99) | 0.604 |
| *A<sub>1</sub> + A<sub>2</sub>* | 3,031 (3.0) | 169 | 1.22 (1.03-1.44) | 0.023 | 150 | 35 | 0.90 (0.60-1.34) | 0.611 |  | 5 | 0.99 (0.39-2.55) | 0.990 |
| *A<sub>2</sub> + O* | 8,714 (8.5) | 470 | 1.16 (1.04-1.29) | 0.008 | 407 | 112 | 1.23 (0.96-1.58) | 0.101 |  | 12 | 0.92 (0.48-1.74) | 0.785 |
| *A<sub>2</sub> + A<sub>2</sub>* | 523 (0.5) | 30 | 1.32 (0.90-1.95) | 0.157 | 28 | 10 | 1.93 (0.87-4.27) | 0.107 |  | 0 | 0 | - |

Legend: N, number of cases, OR, odds ratio; CI, confidence interval; p, p value. Odds ratios were calculated in a multivariable logistic regression model containing information on age, sex, BMI, diabetes mellitus, arterial hypertension, smoking status and month of testing.
\* Infections reported for the months January to July, excluding August and September.

## Discussion

Individual risk assessment for adults aged 18 to 61 years without serious health issues remains uncertain. More information on risk factors for this part of the working population may inform decision making throughout the course of the pandemic. We present data of one of the largest studies in this population which have been conducted so far. In comparison to data for the general population collected with the European Health Interview Survey, our study population showed lower rates of diabetes, arterial hypertension, and nicotine abuse (see supplemental Table S2)^18^. Notably, women are overrepresented in our cohort, which partly reflects the preponderance of women among registered volunteers. The pattern of our results suggests that exposition to the virus in the environment, captured by numbers of infections per month, dominated other risk factors for contracting SARS-CoV-2. Host susceptibility appeared to explain little variation and the bimodal distribution of the percentage of positive tests across age groups (Figure 1 and Table 2) most likely reflected age-group specific exposition patterns and behavior.

Our data are compatible with a protective effect of active smoking to contract SARS-CoV-2 which has been observed in other large studies also^2-3, 12^. Mechanistic explanations have been proposed, e.g. that nicotine might exert anti-inflammatory effects or that nitric oxide inhibits viral replication and invasion of target cells in the respiratory tract^19-20^. However, owing to the character of smoking as sensitive self-reported information we and others cannot exclude a reporting bias. For example, smokers could ask more often for tests driven by mild respiratory symptoms than non-smokers. If so, any inference on the percentage of positive tests by smoking status would be biased.

In line with previous reports and meta analyses, we found that BG A is a significant, albeit weak, risk factor for contracting SARS-CoV-2^10, 12-13^. Notably, the investigation of ABO subgroups did not provide a consistent explanation for this association. Participants with A_1_ versus A_2_ phenotype showed comparable rates of infections. Since the A-epitope density is more than four-fold higher in BG A_1_ versus A_2_, our data therefore argue against the hypothesis that A-epitopes facilitate SARS-CoV-2 entry into cells^14^. Genetic variance in *ABO* genes has been associated with the risk for venous thromboembolism, stroke, coronary artery disease, and acute respiratory distress syndrome regardless of COVID-19^21-23^. The impact of glycosylation patterns on the clearance of plasma proteins (e.g. of the von Willebrand Factor) may result for example in differences in end-stage coagulation and thrombosis^24-25^. This would also explain a negative impact of BG A in high-risk populations but not in low-risk populations like ours^10-11, 26^. In exploratory analyses, BG B was associated with a greater risk of severe respiratory infections and respiratory hospitalization. However, genotype-specific analyses did not reveal a conclusive pattern of associations (Table 3 and Table S2) and BG B has not been described consistently as risk factor for severe COVID-19 courses^10, 12-13^. Therefore, we interpret this association as incidental finding.

Only 3.6% of participants who had been tested positive, reported hospitalizations due to respiratory infections. This rate is substantially lower than the 14% average hospitalization rate for COVID-19 for Germany^7^. Yet, the hospitalization rate of 1.8% among infected crew members of an aircraft carrier with a median age of 28 years was even lower^26^. These apparent discrepancies can partly be explained by differences between the study populations with respect to two major risk factors for respiratory hospitalizations, age, and BMI. Based on multivariable regression modeling, the risk of hospitalization due to RTI was 5.3-fold higher for individuals aged 55-61 years compared to 18-24 years. This impact of age on the risk of hospitalization due to SARS-CoV-2 among 18 to 61-year-old adults is remarkable and appears to be more pronounced than for influenza. The influenza burden by age group changes from season to season, but in contrast to SARS-CoV-2, influenza-associated rates of hospitalizations are typically higher for the very young and the very old^27^. The second major risk factor is BMI. In multivariable regression modeling, overweight (BMI 25 to <30 kg/m^2^) increased the risk of hospitalization (odds ratio 1.35, p=0.048) and obesity class II (BMI 35 to <40 kg/m^2^) approximately doubled the risk of hospitalization (odds ratio 2.1, p=0.01). The prevalence of obesity in a population may therefore be a major factor determining the burden of COVID-19. While our data stand out with respect to the granularity, others reported already that overweight and obesity might be strong risk factors for COVID-19 hospitalization and mortality^3, 12, 28^. Yet, obesity also has a strong impact on the respiratory hospitalizations for influenza and might thus not reflect COVID-19 specific pathology ^29^.

Only 22 out of 7,948 participants (0.28%) needed mechanical ventilation in our cohort. This low number fits to a weighted case fatality ratio of 0.11% calculated for a population with comparable sex- and age-distribution based on national data (Supplemental Table S3). It is also in line with rates of ICU admissions in smaller studies on healthy populations and larger studies on COVID-19-related mortality ^3-4, 26^.

Our study has several limitations: First, we analyzed self-reported data. Test results from oro- or nasopharyngeal swabs were not verified. Further, gender-specific or smoker-specific behavior in response to the pandemic may have biased our data ^30^. Second, the phenotype definitions based on self-reported symptoms without objective measurements. Therefore, we introduced respiratory hospitalization as a more stringent – but non-standard - definition for severity in order to exclude hospitalizations for isolation or social indications. Third, negative cases had no confirmed COVID-19 infection during the past 6 months. However, this did not preclude asymptomatic or mild to moderate SARS-CoV-2 infections, which had not been diagnosed. Forth, rates of positive tests must not be interpreted as incidences because our study population is not a random sample of the general population. Fifth, by design, we could not collect data on events, which incapacitated volunteers to respond. The infection-fatality-rate of adults below 61 years of age is low, but the rate of respiratory hospitalization calculated from self-reported data systematically underestimates the true rate of hospitalizations by this margin.

Taken together, the data show a moderate risk of respiratory hospitalizations due to SARS-CoV-2 infection in this cohort of mostly healthy participants ranging between 18 and 61 years of age. The risk for hospitalization, however, varied substantially depending on age and BMI. Blood group A had no impact on this endpoint. Our results may be used for individual counseling and risk-stratified preventive measures.

## Data Availability

The authors confirm that the data supporting the results of this study are available in the article [and/or] its supplementary materials.

## Acknowledgements

We are grateful to all registered DKMS donors who participated in this study. Further, we would like to acknowledge the dedicated work of many of our coworkers in different departments of DKMS who facilitated this study. Especially, we would like to name Alois Grathwohl, Heike Fischer, Stefanie Diesch, Michaela Jaeger, Rolando Silva, Nina Belser, Sandra Fritschi, Julia Chatelain, Larissa Wolf, Jonas Wolf, and Marcel Eggert (HLA Service), Theodora Fouki, Clemens Fritschi, Philipp Rau-Endrulat, Philipp Krempel, Jochen Schnitzer, Kai Höfler, Julia Pingel and Mike Bretz (IT), Nina Louis, Sonja Krohn, Kay Beutling, and Konstanze Burkard (Corporate Communications), Alessandro Haemmerle, Claus Haibt, and Ralf Neumann (Data Management), and Daniel Schefzyk (Bioinformatics). Finally, we would like to acknowledge a research grant from BMBF (reference number 01KI20177) which in parts facilitated this study.

## Author contributions

A.H. Schmidt, S.N. Bernas, J. Schetelig, and H. Baldauf designed the study.

S. Wendler, F. Heidenreich, J. Schetelig, R. Real, and S.N. Bernas performed systematic literature searches addressing different aspects of design and risk factors for this study.

S. Wendler, S.N. Bernas, R. Real, H. Baldauf, J. Schetelig, and A.H. Schmidt developed the health questionnaire.

C. Bunzel, D. Endert, R. Barth, S.N. Bernas, J. Sauter, and H. Baldauf designed and implemented the data protection concept.

J. Sauter, R. Barth, J. Hofmann, J. Markert, S.N. Bernas, D. Endert designed the database, validated data entry and export, and verified the data download.

H. Baldauf and J. Schetelig performed the statistical analysis.

H. Baldauf, S.N. Bernas, and J. Sauter had access to the underlying data and verified their integrity.

All authors contributed to the interpretation of the data.

J. Schetelig wrote the first draft of the manuscript. All authors reviewed the paper and approved the final version of the manuscript.

## Notes

### Competing Interest Statement

The authors have declared no competing interest.

### Author Declarations

Institutional Review Board of the Technische Universitaet Dresden (IRB00001473)

## References

1. Zhou F, Yu T, Du R, Fan G, Liu Y, Liu Z, et al. Clinical course and risk factors for mortality of adult inpatients with COVID-19 in Wuhan, China: a retrospective cohort study. Lancet. 2020 Mar 11.

2. Petrilli CM, Jones SA, Yang J, Rajagopalan H, O’Donnell L, Chernyak Y, et al. Factors associated with hospital admission and critical illness among 5279 people with coronavirus disease 2019 in New York City: prospective cohort study. BMJ. 2020 May 22;369:m1966.

3. Williamson EJ, Walker AJ, Bhaskaran K, Bacon S, Bates C, Morton CE, et al. Factors associated with COVID-19-related death using OpenSAFELY. Nature. 2020 Aug;584(7821):430–6.

4. Karagiannidis C, Mostert C, Hentschker C, Voshaar T, Malzahn J, Schillinger G, et al. Case characteristics, resource use, and outcomes of 10?021 patients with COVID-19 admitted to 920 German hospitals: an observational study. Lancet Respir Med. 2020 Sep;8(9):853–62.

5. Ryan C, Minc A, Caceres J, Balsalobre A, Dixit A, Ng BK, et al. Predicting severe outcomes in Covid-19 related illness using only patient demographics, comorbidities and symptoms. Am J Emerg Med. 2020 Sep 9.

6. Boehmer TK, DeVies J, Caruso E, van Santen KL, Tang S, Black CL, et al. Changing Age Distribution of the COVID-19 Pandemic - United States, May-August 2020. MMWR Morb Mortal Wkly Rep. 2020 Oct 2;69(39):1404–9.

7. Coronavirus Disease 2019 (COVID-19) Daily Situation Report of the Robert Koch Institute, October 13, 2020. [cited 18.10.2020]; Available from: https://www.rki.de/EN/Content/infections/epidemiology/outbreaks/COVID-19/Situationsberichte_Tab.html

8. Schmidt AH, Sauter J, Baier DM, Daiss J, Keller A, Klussmeier A, et al. Immunogenetics in stem cell donor registry work: The DKMS example (Part 1). Int J Immunogenet. 2020 Feb;47(1):13–23.

9. Schmidt AH, Mengling T, Hernandez-Frederick CJ, Rall G, Pingel J, Schetelig J, et al. Retrospective Analysis of 37,287 Observation Years after Peripheral Blood Stem Cell Donation. Biol Blood Marrow Transplant. 2017 Jun;23(6):1011–20.

10. Ellinghaus D, Degenhardt F, Bujanda L, Buti M, Albillos A, Invernizzi P, et al. Genomewide Association Study of Severe Covid-19 with Respiratory Failure. N Engl J Med. 2020 Jun 17.

11. Barnkob MB, Pottegård A, Støvring H, Haunstrup TM, Homburg K, Larsen R, et al. Reduced prevalence of SARS-CoV-2 infection in ABO blood group O. Blood Adv. 2020 Oct 27;4(20):4990–3.

12. Shelton JF, Shastri AJ, Ye C, Weldon CH, Filshtein-Somnez T, Coker D, et al. Trans-ethnic analysis reveals genetic and non-genetic associations with COVID-19 susceptibility and severity. 2020.

13. Golinelli D, Boetto E, Maietti E, Fantini MP. The association between ABO blood group and SARS-CoV-2 infection: A meta-analysis. PLoS One. 2020;15(9):e0239508.

14. Cooling L. Technical Manual 18th Edition. In: Fung MK, Grossmann BJ, Hillyer CD, Westhoff CM, editors. Technical Manual. Bethesda: American Association of Blood Banks (ABB); 2014.

15. Lange V, Bohme I, Hofmann J, Lang K, Sauter J, Schone B, et al. Cost-efficient high-throughput HLA typing by MiSeq amplicon sequencing. BMC Genomics. 2014 Jan 24;15:63.

16. Lang K, Wagner I, Schöne B, Schöfl G, Birkner K, Hofmann JA, et al. ABO allele-level frequency estimation based on population-scale genotyping by next generation sequencing. BMC Genomics. 2016 May 20;17:374.

17. Coronavirus Disease 2019 (COVID-19) Daily Situation Report of the Robert Koch Institute 07.10.2020. [cited 18.10.2020]; Available from: https://www.rki.de/EN/Content/infections/epidemiology/outbreaks/COVID-19/Situationsberichte_Tab.html

18. Saß A-C, Lange C, Finger JD, Allen J, Born S, Hoebel J, et al. “Gesundheit in Deutschland aktuell” - Neue Daten für Deutschland und Europa Hintergrund und Studienmethodik von GEDA 2014/2015- EHIS. Journal of Health Monitoring. 2017;2(1):83–90.

19. Farsalinos K, Eliopoulos E, Leonidas DD, Papadopoulos GE, Tzartos S, Poulas K. Nicotinic Cholinergic System and COVID-19: In Silico Identification of an Interaction between SARS-CoV-2 and Nicotinic Receptors with Potential Therapeutic Targeting Implications. Int J Mol Sci. 2020 Aug 13;21(16).

20. Usman MS, Siddiqi TJ, Khan MS, Patel UK, Shahid I, Ahmed J, et al. Is there a smoker’s paradox in COVID-19? BMJ Evid Based Med. 2020 Aug 11.

21. Dentali F, Sironi AP, Ageno W, Turato S, Bonfanti C, Frattini F, et al. Non-O blood type is the commonest genetic risk factor for VTE: results from a meta-analysis of the literature. Semin Thromb Hemost. 2012 Jul;38(5):535–48.

22. Dichgans M, Malik R, König IR, Rosand J, Clarke R, Gretarsdottir S, et al. Shared genetic susceptibility to ischemic stroke and coronary artery disease: a genome-wide analysis of common variants. Stroke. 2014 Jan;45(1):24–36.

23. Reilly JP, Meyer NJ, Shashaty MG, Anderson BJ, Ittner C, Dunn TG, et al. The ABO Histo-Blood Group, endothelial activation, and acute respiratory distress syndrome risk in critical illness. J Clin Invest. 2020 Sep 15.

24. Song J, Chen F, Campos M, Bolgiano D, Houck K, Chambless LE, et al. Quantitative Influence of ABO Blood Groups on Factor VIII and Its Ratio to von Willebrand Factor, Novel Observations from an ARIC Study of 11,673 Subjects. PLoS One. 2015;10(8):e0132626.

25. Suhre K, Arnold M, Bhagwat AM, Cotton RJ, Engelke R, Raffler J, et al. Connecting genetic risk to disease end points through the human blood plasma proteome. Nat Commun. 2017 Feb 27;8:14357.

26. Boudin L, Janvier F, Bylicki O, Dutasta F. ABO blood groups are not associated with risk of acquiring the SARS-CoV-2 infection in young adults. Haematologica. 2020 Jul 23.

27. Centers for Disease Control and Prevention: Past Seasons Estimated Influenza Disease Burden. [cited 18.10.2020]; Available from: https://www.cdc.gov/flu/about/burden/past-seasons.html

28. Popkin BM, D. S, Green WD, Beck MA, Algaith T, Herbst CH, et al. Individuals with obesity and COVID-19: A global perspective on the epidemiology and biological relationships. Obes Rev. 2020 Nov;21(11):e13128.

29. Kwong JC, Campitelli MA, Rosella LC. Obesity and respiratory hospitalizations during influenza seasons in Ontario, Canada: a cohort study. Clin Infect Dis. 2011 Sep;53(5):413–21.

30. Mindell JS, Giampaoli S, Goesswald A, Kamtsiuris P, Mann C, Männistö S, et al. Sample selection, recruitment and participation rates in health examination surveys in Europe--experience from seven national surveys. BMC Med Res Methodol. 2015 Oct 5;15:78.

