## Supplemental Material for "Severity of Respiratory Infections due to SARS-CoV-2 in Working Population: Age and Body Mass Index Outweigh ABO Blood Group"

### Supplemental Material 1. Translated and annotated questionnaire.

| | |
|---|---|
| 1 | Have you been tested for coronavirus infection? ["Yes", "No"] |

↓ **Yes** — **No → END**

| | |
|---|---|
| 2 | Was the test performed by a throat/nasal swab? ["Yes", "No"] |

↓ **Yes** — **No → END**

| | |
|---|---|
| 3 | In which month was the last test for coronavirus performed? [Dec, Jan, Feb, Mar, Apr, May, Jun, Jul, Aug, Sep] |
| 4 | Did you take any antihypertensive medication in 2019? ["Yes", "No"] |
| 5 | Did you take any Diabetes medication in 2019? ["Yes", "No"] |
| 6 | Did you smoke regularly in 2019? ["No, I have never smoked.", "No, but I used to smoke regularly (before 2019).", "Yes, I smoked regularly in 2019." ] |
| 7 | Please enter your height in cm. |
| 8 | Please enter your weight in kg. |

| | |
|---|---|
| 9 | Has SARS-CoV-2 (COVID-19) been detected at least once? [ "Yes", "No", "I have not yet received the test result."] |

↓ **Yes** — **No / not yet → END**

| | |
|---|---|
| 10 | Did you experience symptoms due to coronavirus infection? ["Yes", "No"] |

↓ **Yes** — **No → END**

| | |
|---|---|
| 11 | Which of the following symptoms did you experience?<br>• shortness of breath ["Yes", "No"]<br>• fever (body temperature above 38.5°C) ["Yes", "No"]<br>• cough ["Yes", "No"]<br>• muscle or body aches ["Yes", "No"]<br>• sore throat ["Yes", "No"]<br>• loss of taste and/or smell ["Yes", "No"]<br>• other symptoms ["Yes", "No"] |

| | |
|---|---|
| 12 | Have you been hospitalized for the coronavirus infection? ["Yes", "No"] |

↓ **Yes** — **No → END**

| | |
|---|---|
| 13 | Did you receive supplemental oxygen (e.g. through a nasal cannula)? ["Yes", "No"] |
| 14 | Did you receive general anesthesia and mechanical ventilation? ["Yes", "No"] |

**END**

Answers were submitted after pressing a confirmation button.

- Questions with a bold frame were used to filter respondents.

- Questions with a light frame did not serve to filter participants.
- Square brackets indicate answer categories.
- All questions needed to be completed before submission.

### Supplemental Figure S1. Flow chart

The flow chart below displays exclusions of participants and the size of the final analysis sets. The questionnaire was successfully mailed to 4,440,895 registered DKMS donors and returned by 924,685 participants. 25 participants subsequently withdrew consent. Their data were deleted.

Additionally, data from 103 participants were excluded from data analysis for a variety of reasons: 64 participants reported a BMI of less than 10 kg/m² or more than 65 kg/m², 29 participants reported a test in December 2019, and 10 participants provided missing or inconsistent answers on testing status and disease symptoms.

Of the remaining 924,557 participants, 748,613 had never been tested for SARS-CoV-2. Participants who reported positive results from other tests (N=15,272), e.g. antibody tests, were excluded. In addition, 3,132 respondents did not know the results of their swab when they answered to the survey. The full analysis set thus contained 157,544 participants.

We defined two analysis populations:

The population to analyze the risk of contracting SARS-CoV-2 comprised 157,544 participants, 7,948 participants who had been tested positive and 149,596 individuals who had been tested negative.

The population to analyze risk factors for COVID-19 severity included all participants who had been tested positive, minus individuals who reported positive tests in August or September (N=1,029) because we could not assume that these participants had already recovered when they completed the health questionnaire and thus were not able to report maximum severity. This population comprised 6,919 participants.

Information on ABO blood group and *ABO* allele-level was available for 79% and 65% of all participants, respectively. The numbers of participants with ABO information are provided in the flow chart below.

Supplemental Figure S1. Flow chart.

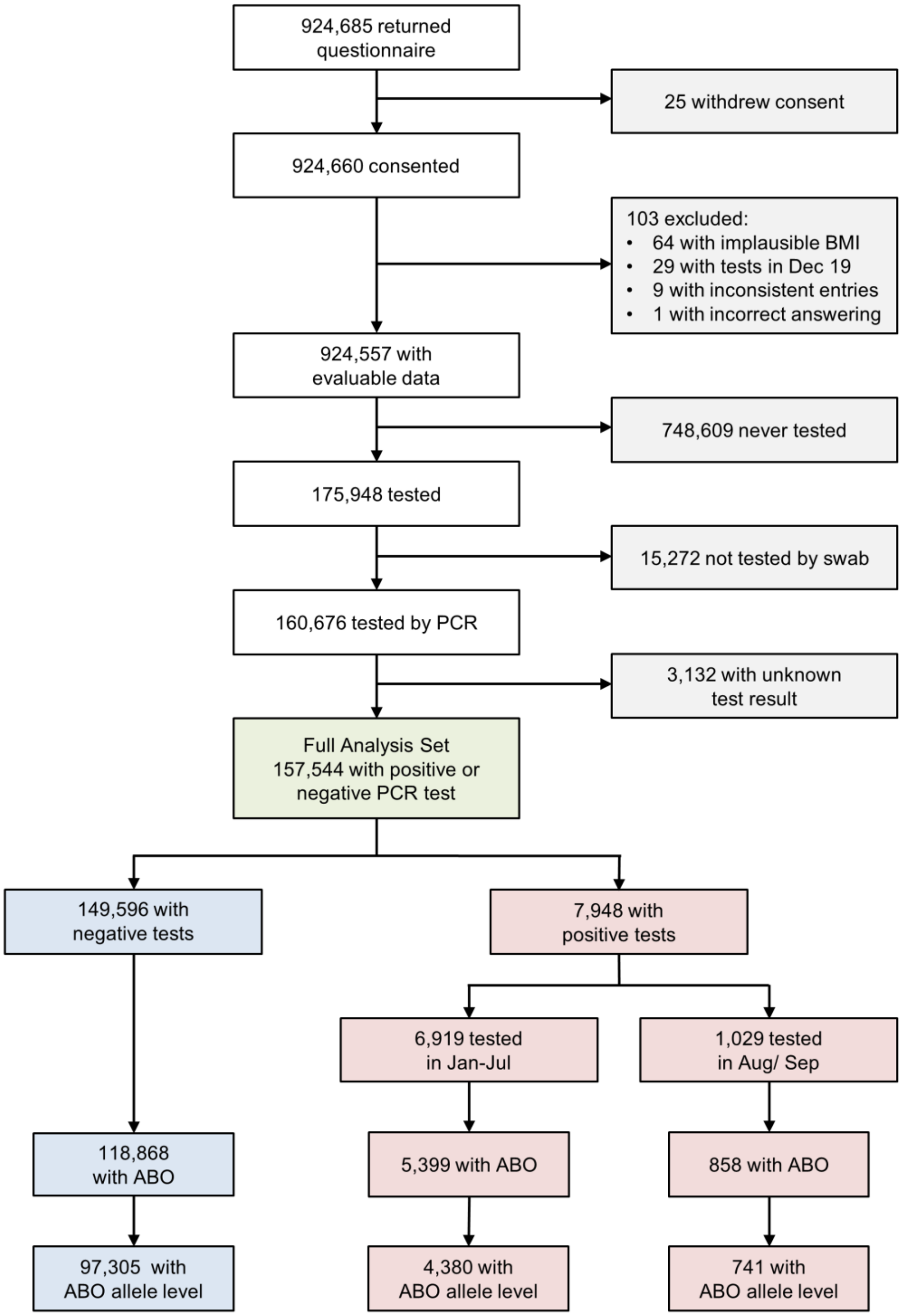

### Supplemental Figure S2. Regional distribution of reported cases

This figure shows maps displaying the regional rate of cases per participants (left) to the regional incidence of SARS-CoV-2 infections as provided by the Robert-Koch-Institute (RKI) (right). In contrast to our study, RKI data cover the whole German population. As displayed below, the study data mirror regional differences in the rate of SARS-CoV-2 positive tests. Regional rates of positive tests ranged from 198.5 to 2713.7 per 100,000 participants (median 780.4) in our study. In comparison, regional incidences of SARS-CoV-2 infections reported by the RKI varied between 52.9 and 731.3 cases per 100,000 inhabitants (median 284.8).

### Supplemental Figure S2. Regional distribution of reported cases.

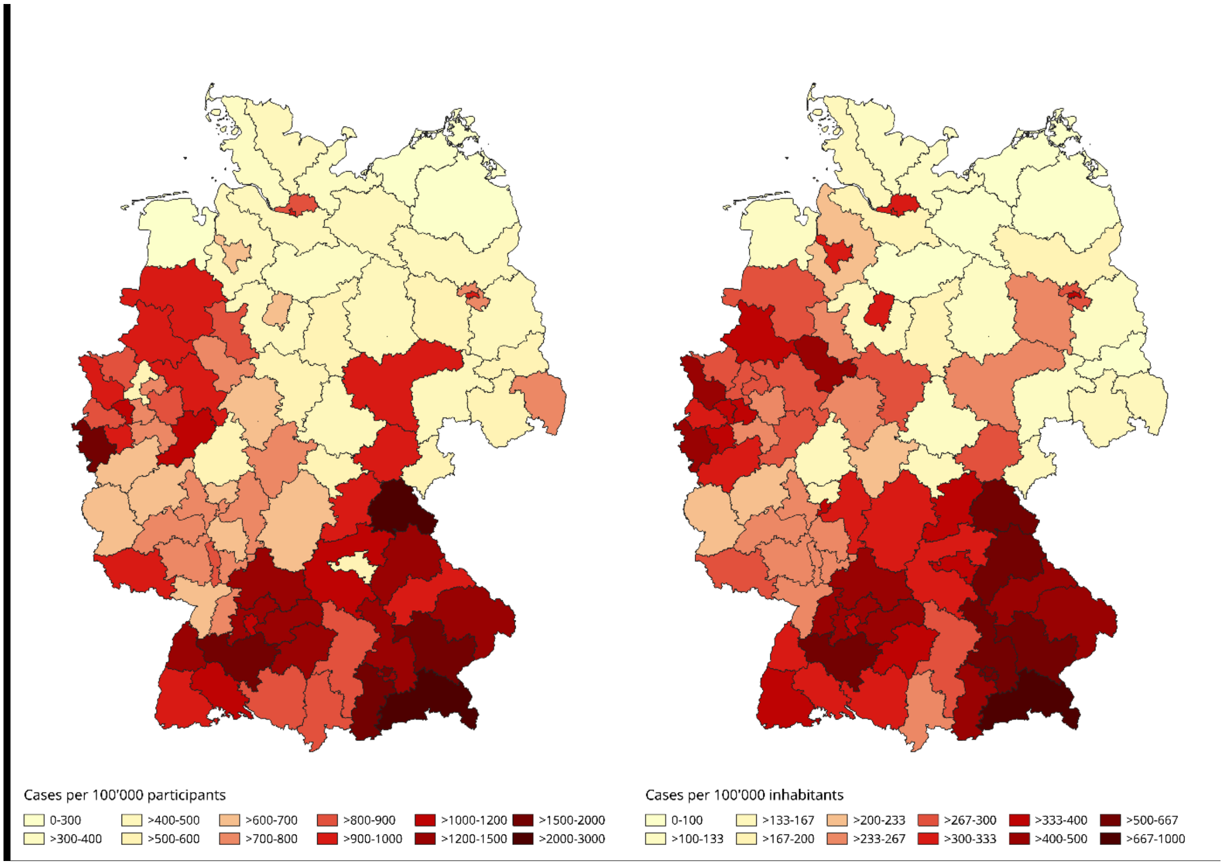


Incidences of SARS-CoV-2 infections in the right panel were reported for the time between January 1st and August 31st, 2020. To calculate the incidence of infections for the whole German population, we downloaded data from the Robert Koch Institute (RKI) as of October 10th, 2020 (https://www.arcgis.com/home/item.html?id=f10774f1c63e40168479a1feb6c7ca74). We mapped the first two digits of German Postal Codes to the regions provided in RKI file according to data downloaded from https://www.suche-postleitzahl.org/downloads. We created the maps with Open Source Software Quantum GIS, version 3.16.0, and Gnu Image Manipulation Program, version 2.8.22.

### Supplemental Table S1. Risk of infection, severe respiratory tract infection, and respiratory hospitalization by *ABO* genotype including rare variants and Rhesus genotype

Analyses on minor *ABO* subgroups were part of exploratory studies to investigate genetic linkage of phenotypes with certain genotypes. These tests were not motivated by biological hypotheses. As a consequence, p-values of these tests have to be interpreted in the context of multiple testing.

Rhesus proteins are expressed only on red blood cells and their precursors. Therefore, we formulated no biological hypothesis on a linkage with SARS-CoV-2 infections. However, conflicting results on associations of rhesus polymorphisms have been reported in large studies (Zietz et al 2020, Niles et al 2020, Latz et al 2020, and Shelton et al 2020 – references provided below). Since we could not rule out confounding through genetic linkage, we checked for associations of the rhesus blood group with the risk of infection and the severity of COVID-19. These tests were also performed in the framework of exploratory testing. Our data did not show a significant association of rhesus gene polymorphism with the respective phenotypes (see Table below).

### References


1. Zietz M, Tatonetti NP. Testing the association between blood type and COVID-19 infection, intubation, and death. medRxiv. 2020 Apr 11.
2. Niles JK, Karnes HE, Dlott JS, Kaufman HW. Association of ABO/Rh with SARS-CoV-2 positivity: The role of race and ethnicity in a female cohort. Am J Hematol. 2020 Oct 16.
3. Latz CA, DeCarlo C, Boitano L, Png CYM, Patell R, Conrad MF, et al. Blood type and outcomes in patients with COVID-19. Ann Hematol. 2020 Sep;99(9):2113-8.
4. Shelton JF, Shastri AJ, Ye C, Weldon CH, Filshtein-Somnez T, Coker D, et al. Trans-ethnic analysis reveals genetic and non-genetic associations with COVID-19 susceptibility and severity. 2020.

**Supplemental Table S1. Risk of infection, severe respiratory tract infection, and respiratory hospitalization by *ABO* genotype including rare variants and Rhesus genotype.**

| Genotype | Contracting SARS-CoV-2 | | | | Evaluable* infections | Severe respiratory tract infections | | | Respiratory hospitalizations | | |
|---|---|---|---|---|---|---|---|---|---|---|---|
| | Tests (%) | Cases | OR (95%-CI) | p | N | N | OR (95%-CI) | p | N | OR (95%-CI) | p |
| $O_1+O_2$ | 18,014 (17.6) | 811 | 1 | | 693 | 152 | 1 | | 21 | 1 | |
| $A_1+O_1$ | 16,386 (16.0) | 884 | 1.20 (1.08-1.32) | 0.001 | 737 | 188 | 1.23 (0.96-1.58) | 0.098 | 17 | 0.78 (0.40-1.50) | 0.452 |
| $O_1+O_1$ | 14,680 (14.3) | 709 | 1.09 (0.98-1.21) | 0.115 | 616 | 156 | 1.20 (0.92-1.55) | 0.176 | 23 | 1.22 (0.66-2.23) | 0.531 |
| $A_1+O_2$ | 9,866 (9.6) | 519 | 1.18 (1.05-1.33) | 0.006 | 442 | 117 | 1.27 (0.96-1.68) | 0.095 | 19 | 1.41 (0.75-2.68) | 0.289 |
| $B_1+O_1$ | 6,907 (6.7) | 325 | 1.07 (0.93-1.23) | 0.340 | 267 | 75 | 1.38 (1.00-1.92) | 0.053 | 16 | 2.07 (1.05-4.08) | 0.035 |
| $O_2+O_2$ | 5,413 (5.3) | 261 | 1.06 (0.92-1.23) | 0.416 | 227 | 61 | 1.28 (0.90-1.82) | 0.162 | 7 | 0.99 (0.41-2.39) | 0.988 |
| $A_2+O_1$ | 5,262 (5.1) | 288 | 1.23 (1.07-1.42) | 0.004 | 246 | 70 | 1.46 (1.04-2.04) | 0.027 | 7 | 0.97 (0.40-2.34) | 0.953 |
| $A_1+A_1$ | 4,612 (4.5) | 214 | 1.03 (0.88-1.21) | 0.735 | 187 | 54 | 1.42 (0.98-2.06) | 0.062 | 5 | 0.85 (0.32-2.31) | 0.755 |
| $A_1+B_1$ | 4,018 (3.9) | 214 | 1.23 (1.05-1.45) | 0.011 | 185 | 53 | 1.47 (1.01-2.13) | 0.042 | 6 | 1.10 (0.44-2.80) | 0.834 |
| $B_1+O_2$ | 3,600 (3.5) | 180 | 1.13 (0.95-1.34) | 0.176 | 151 | 56 | 2.17 (1.48-3.19) | <.001 | 13 | 3.05 (1.47-6.32) | 0.003 |
| $A_2+O_2$ | 3,119 (3.0) | 163 | 1.15 (0.96-1.38) | 0.120 | 145 | 38 | 1.30 (0.85-1.97) | 0.224 | 5 | 1.17 (0.43-3.20) | 0.757 |
| $A_1+A_2$ | 3,031 (3.0) | 169 | 1.27 (1.06-1.51) | 0.009 | 150 | 35 | 1.01 (0.66-1.55) | 0.951 | 5 | 1.09 (0.40-2.96) | 0.872 |
| $A_2+B_1$ | 1,278 (1.2) | 69 | 1.34 (1.03-1.75) | 0.029 | 59 | 17 | 1.46 (0.80-2.66) | 0.215 | 0 | 0 - | - |
| $O_1+O_3$ | 1,228 (1.2) | 68 | 1.12 (0.86-1.46) | 0.412 | 61 | 18 | 1.54 (0.86-2.77) | 0.147 | 4 | 2.28 (0.74-7.03) | 0.150 |
| $B_1+B_1$ | 835 (0.8) | 37 | 1.04 (0.74-1.48) | 0.806 | 33 | 6 | 0.80 (0.32-2.02) | 0.642 | 1 | 0.85 (0.11-6.80) | 0.882 |
| $O_2+O_3$ | 749 (0.7) | 32 | 1.02 (0.70-1.48) | 0.928 | 25 | 5 | 1.00 (0.36-2.72) | 0.993 | 0 | 0 - | - |
| $A_1+O_3$ | 690 (0.7) | 41 | 1.35 (0.96-1.90) | 0.080 | 35 | 9 | 1.12 (0.51-2.49) | 0.777 | 0 | 0 - | - |
| $A_2+A_2$ | 523 (0.5) | 30 | 1.38 (0.93-2.04) | 0.108 | 28 | 10 | 2.17 (0.96-4.87) | 0.061 | 0 | 0 - | - |
| $B_1+O_3$ | 243 (0.2) | 8 | 0.66 (0.32-1.37) | 0.264 | 6 | 2 | 1.77 (0.32-9.92) | 0.518 | 0 | 0 - | - |
| $A_2+O_3$ | 240 (0.2) | 13 | 1.19 (0.66-2.14) | 0.553 | 11 | 4 | 1.75 (0.50-6.12) | 0.384 | 0 | 0 - | - |
| $O_1+O_9$ | 162 (0.2) | 5 | 0.66 (0.26-1.65) | 0.377 | 4 | 1 | 1.27 (0.12-12.9) | 0.841 | 0 | 0 - | - |
| $O_1+O_{49}$ | 123 (0.1) | 7 | 1.41 (0.63-3.13) | 0.405 | 6 | 2 | 1.62 (0.29-9.11) | 0.586 | 0 | 0 - | - |
| $O_1+O_{48}$ | 121 (0.1) | 2 | 0.40 (0.10-1.68) | 0.213 | 2 | 0 | 0 - | - | 0 | 0 - | - |
| $O_2+O_9$ | 115 (0.1) | 7 | 1.23 (0.55-2.74) | 0.614 | 6 | 1 | 0.74 (0.08-6.54) | 0.787 | 0 | 0 - | - |
| $A_1+O_9$ | 108 (0.1) | 9 | 2.28 (1.10-4.73) | 0.027 | 7 | 2 | 1.23 (0.23-6.56) | 0.806 | 0 | 0 - | - |
| $O_1+O_{50}$ | 98 (0.1) | 6 | 1.29 (0.54-3.07) | 0.569 | 6 | 0 | 0 - | - | 0 | 0 - | - |
| $O_2+O_{49}$ | 91 (0.1) | 2 | 0.55 (0.13-2.29) | 0.410 | 2 | 0 | 0 - | - | 0 | 0 - | - |
| $A_1+O_{49}$ | 84 (0.1) | 5 | 1.38 (0.54-3.55) | 0.506 | 5 | 0 | 0 - | - | 0 | 0 - | - |
| $O_2+O_{50}$ | 83 (0.1) | 4 | 0.97 (0.34-2.75) | 0.951 | 4 | 1 | 1.50 (0.15-14.9) | 0.732 | 0 | 0 - | - |
| $A_1+O_{48}$ | 69 (0.1) | 1 | 0.35 (0.05-2.60) | 0.307 | 1 | 0 | 0 - | - | 0 | 0 - | - |
| Other | 678 (0.7) | 38 | 1.16 (0.82-1.64) | 0.403 | 33 | 7 | 0.97 (0.41-2.29) | 0.938 | 1 | 1.10 (0.14-8.50) | 0.928 |
| **Rhesus** | | | | | | | | | | | |
| Rh(D)+ | 103,444 (82.6) | 5,207 | 1 | | 4,504 | 1,189 | 1 | | 157 | 1 | |
| Rh(D)- | 21,738 (17.4) | 1,053 | 0.96 (0.89-1.03) | 0.243 | 898 | 222 | 0.91 (0.77-1.08) | 0.277 | 41 | 1.34 (0.94-1.91) | 0.110 |

Legend: N, number of cases, OR, odds ratio; CI, confidence interval; p, p value. Odds ratios are adjusted for age, sex, BMI, diabetes mellitus, arterial hypertension, smoking status and month of testing. * Infections reported for the months January to July, excluding August and September.

### Supplemental Table S2. Comparison of study population and general population by key health status characteristics

To relate the overall health status of the study sample to the German general population, we used data from the latest German health update (GEDA 2014/2015; Saß et al., 2017) to compare 12-month prevalence rates of diabetes (Heidemann et al., 2017), hypertonia (Neuhauser et al., 2017), BMI (Schienkiewitz et al., 2017), and smoking behavior (Zeiher & Kuntz, 2017). Comparisons are based on aggregated data from GEDA respondents between 18 and less than 65 years of age. Data from the GEDA study and English translations of GEDA questionnaire items were downloaded from gbe-bund.de on October 8th, 2020. The questionnaire for our study was not designed with the aim of direct comparisons with the GEDA data.

Of note, the GEDA age group of 45-65-year-old participants included more older participants, as for our study the upper age limit was 61 years, defined by the maximum age for unrelated volunteer stem cell donation. This raises the possibility that age effects in the 45-65 age group are due to an underrepresentation of older participants in our study.

Supplemental Table S2 lists the prevalence of diabetes mellitus, the prevalence of arterial hypertension, the distribution of the Body Mass Index and the distribution of non-smokers among participants of this study and the GEDA study.

Comments:

The prevalence of diabetes mellitus in our study is comparable to the general population for participants aged below 45 years but a lower prevalence of diabetes among our participants aged 45 to 60 years is likely. The GEDA study recorded diabetes status via a single question: “Did you have one of the following diseases or health problems in the last 12 months?”. The list of possible answer categories included the option “diabetes (not gestational diabetes)”. Our questionnaire recorded diabetes status also via a single question: “Did you take any Diabetes medication in 2019?” (answer categories: “Yes”, “No”). Diabetes rates reported in our sample and the GEDA study are, thus, not strictly comparable. Our indicator is likely stricter than the GEDA indicator since medication is not the only available treatment option and only about 73% of German patients (45-79 years) with known diabetes receive medication (Robert Koch-Institut, 2019).

In comparison to GEDA participants, our participants showed consistently lower rates of arterial hypertension in all age groups and both sexes. The GEDA study recorded high blood pressure status using three questions (answer categories: “Yes”, “No”). A positive answer to "Have you ever been diagnosed with high blood pressure by a doctor?" and at least one positive answer to one of the two questions "Did you have high blood pressure in the last 12 months?" or "Are you currently taking antihypertensive medicine?" are taken as an indicator of high blood pressure in the last 12 months. Our questionnaire recorded blood pressure status via a single question: “Did you take any antihypertensive medication in 2019?” (answer categories: “Yes”, “No”). Our indicator was defined stricter than the GEDA indicator, as only about 88% of German patients with known hypertension receive antihypertensive medication intervention. In addition, medication rates show strong variation by age (higher in older patients) and sex (higher in women; Neuhauser et al., 2015).

Across all age groups and both sexes the distribution of BMI categories is comparable between our and GEDA participants. The GEDA study recorded respondents’ height and weight via two questions: “What is your height without shoes?” (cm) and “How much do you weigh without clothing and shoes?” (kg). Pregnant women were asked to report their weight before pregnancy. Our questionnaire elicited height and weight via two similar questions: “Please enter your height in cm” and “Please enter your weight in kg”. Although our questions did not include instructions on how height and weight were to be measured and did not mention pregnancy, results are likely well comparable.

Across all age groups and both sexes, the proportion of never-smokers is consistently higher in our sample in comparison to the GEDA sample. The GEDA study recorded smoking status with a single question: "Do you smoke?" (answer categories: "yes, daily", "yes, occasionally", "no, no longer", "have never smoked") to differentiate between current smokers (daily or occasionally), former smokers, and never-smokers. Our questionnaire recorded smoking status with a single question “Did you smoke regularly in 2019?” (answer categories: “No, I have never smoked”, “No, but I used to smoke before 2019”, and “Yes, I smoked regularly in 2019”). As not all answer categories are strictly comparable, the comparison was restricted to never-smokers.

### References


Heidemann, C., Kuhnert, R., Born, S., & Scheidt-Nave, C. (2017). 12-Monats-Prävalenz des bekannten Diabetes mellitus in Deutschland. *Journal of Health Monitoring*, *2*. Robert Koch-Institut, Epidemiologie und Gesundheitsberichterstattung. https://doi.org/10.17886/RKI-GBE-2017-008

Neuhauser, H. K., Adler, C., Rosario, A. S., Diederichs, C., & Ellert, U. (2015). Hypertension prevalence, awareness, treatment and control in Germany 1998 and 2008–11. Journal of Human Hypertension, 29(4), 247–253. https://doi.org/10.1038/jhh.2014.82

Neuhauser, H., Kuhnert, R., & Born, S. (2017). 12-Monats-Prävalenz von Bluthochdruck in Deutschland. *Journal of Health Monitoring (Bd. 2). Robert Koch-Institut, Epidemiologie und Gesundheitsberichterstattung*. https://doi.org/10.17886/RKI-GBE-2017-007

Robert Koch-Institut (2017). Fragebogen zur Studie „Gesundheit in Deutschland aktuell“ GEDA 2014/2015-EHIS. *Journal of Health Monitoring*, *2*. https://doi.org/10.17886/RKI-GBE-2017-014

Robert Koch-Institut (2019). *Diabetes in Deutschland*. https://doi.org/10.25646/6284

Saß, A.-C., Lange, C., Finger, J. D., Allen, J., Born, S., Hoebel, J., Kuhnert, R., Müters, S., Thelen, J., Schmich, P., Varga, M., Lippe, E. von der, Wetzstein, M., & Ziese, T. (2017). „Gesundheit in Deutschland aktuell“– Neue Daten für Deutschland und Europa Hintergrund und Studienmethodik von GEDA 2014/2015-EHIS. *Journal of Health Monitoring*, *2*. https://doi.org/10.17886/RKI-GBE-2017-012

Schienkiewitz, A., Mensink, G., Kuhnert, R., & Lange, C. (2017). Übergewicht und Adipositas bei Erwachsenen in Deutschland. *Journal of Health Monitoring*. https://doi.org/10.17886/RKI-GBE-2017-025

**Supplemental Table S2. Comparison of study population and general population by key health status characteristics.**

| Characteristic | Study | Percentages of individuals with this characteristic (absolute numbers for DKMS study) | | | | | | |
|---|---|---|---|---|---|---|---|---|
| | | **18-29 years, Female** | **18-29 years, Male** | **30-44 years, Female** | **30-44 years, Male** | **45-65 years, Female** | **45-65 years, Male** | **Total (18-65ys)** |
| Diabetes | DKMS | 1.8 (662) | 1.3 (157) | 2.1 (1,033) | 1.6 (337) | 2.1 (517) | 3.0 (398) | 2.0 (3,104) |
| | GEDA | 1.1 | 0.5 | 1.4 | 2.0 | 5.2 | 9.4 | 4.3 |
| Hypertension | DKMS | 2.5 (919) | 3.0 (350) | 4.9 (2,415) | 8.0 (1,734) | 17.1 (4,156) | 24.4 (3,273) | 8.2 (12,847) |
| | GEDA | 4.2 | 4.4 | 9.0 | 14.5 | 31.6 | 38.3 | 21.3 |
| BMI <18.5 | DKMS | 3.6 (1,355) | 1.9 (230) | 1.7 (833) | 0.3 (57) | 1.1 (273) | 0.1 (12) | 1.8 (2,760) |
| | GEDA | 7.5 | 3.2 | 2.4 | 0.1 | 1.6 | 0.2 | 2.0 |
| BMI 18.5-<25 | DKMS | 66.5 (24,855) | 60.0 (7,098) | 56.4 (27,585) | 42.2 (9,169) | 50.0 (12,160) | 32.2 (4,318) | 54.1 (85,185) |
| | GEDA | 66.4 | 62.8 | 56.1 | 39.9 | 48.3 | 29.7 | 47.4 |
| BMI 25-<30 | DKMS | 19.2 (7,184) | 28.6 (3,387) | 25.1 (12,301) | 41.1 (8,931) | 29.8 (7,233) | 47.0 (6,308) | 28.8 (45,344) |
| | GEDA | 16.5 | 25.1 | 24.2 | 42.6 | 30.5 | 48.2 | 33.5 |
| BMI ≥ 30 | DKMS | 10.7 (3,983) | 9.4 (1,114) | 16.8 (8,205) | 16.4 (3,550) | 19.1 (4,631) | 20.7 (2,772) | 15.4 (24,255) |
| | GEDA | 9.7 | 8.9 | 17.3 | 17.3 | 19.6 | 21.9 | 17.2 |
| Never smoked | DKMS | 70.0 (26,147) | 67.3 (7,959) | 52.0 (25,422) | 52.5 (11,389) | 53.4 (12,979) | 51.3 (6,886) | 57.6 (90,782) |
| | GEDA | 55.7 | 52.6 | 46.1 | 36.0 | 43.4 | 33.8 | 42.8 |

Legend: BMI, body mass index in $kg/m^2$

**Supplemental Table S3. Number of SARS-CoV-2 infections and case fatality rate based on national epidemiological data in younger individuals**

Mortality could not be derived from our data. Instead, we extracted information on age-and gender-specific COVID-19 related mortality from population tables for Germany provided by the RKI (see Table below). Based on these figures, the weighted case-fatality ratio for our study population was 0.11% given the preponderance of young female adults. This would translate into 17 COVID-19 related deaths among the 4,440,895 contacted volunteers.

**Supplemental Table S3. Number of SARS-CoV-2 infections and case fatality rate based on national epidemiological data in younger individuals.**

| Sex/age group | Number of contacted volunteers in DKMS study | Incidence of SARS-CoV-2 infections*$ | Case fatality rate$ |
|---|---|---|---|
| **Female, 18 – 34 years** | 1,437,622 | 0.383% | 0.010% |
| **Female, 35 – 61 years** | 1,280,151 | 0.349% | 0.126% |
| **Male, 18 – 34 years** | 799,567 | 0.379% | 0.018% |
| **Male, 35 – 61 years** | 923,555 | 0.332% | 0.330% |

* The cumulative incidence of SARS-CoV-2 infections is reported for the time period between January 1st and August 31st, 2020.
$ To calculate the incidence of infections and the case fatality rate for the German population we downloaded Data from the Robert Koch Institute (RKI) as of January 11th, 2021 (https://www.arcgis.com/home/item.html?id=f10774f1c63e40168479a1feb6c7ca74). Population sizes by sex and age group for the German population as of December 31st, 2019 were downloaded from German Federal Statistical Office (Destatis). (Table 12411-0006 on 02.10.2020 (https://www-genesis.destatis.de/genesis/online).
Comment: The RKI reported infections for age groups but not the exact age of affected individuals. Individuals ranging between 15 to 17 years of age are not represented in our cohort. Therefore, we used RKI numbers for the age group ranging from 15 to 34 years as approximation to estimate incidences and the case fatality rate for our subgroup of 18 to 34 year old participants. Further, we used RKI numbers for the age group ranging from 35 to 59 years to estimate incidences and the case fatality rate for our subgroup of 35 to 61 year old participants.
